# Identifying high-risk comorbidities of short and long-term opioid prescription use

**DOI:** 10.1101/2021.11.23.21266717

**Authors:** Mariela V Jennings, Hyunjoon Lee, Daniel B Rocha, Sevim B Bianchi, Brandon J Coombes, Richard C Crist, Annika Faucon, Yirui Hu, Rachel L Kember, Travis T Mallard, Maria Niarchou, Melissa N Poulsen, Peter Straub, Richard D Urman, Colin G Walsh, PsycheMERGE Substance Use Disorder Workgroup, Lea K Davis, Jordan W Smoller, Vanessa Troiani, Sandra Sanchez-Roige

**Affiliations:** Department of Psychiatry, University of California San Diego; Psychiatric and Neurodevelopmental Genetics Unit, Center for Genomic Medicine, Massachusetts General Hospital; Phenomic Analytics and Clinical Data Core, Geisinger; Department of Quantitative Health Sciences, Mayo Clinic; Department of Psychiatry, University of Pennsylvania Perelman School of Medicine; Vanderbilt Genetics Institute, Vanderbilt University Medical Center; Department of Population Health Sciences, Geisinger; Department of Psychiatry, University of Pennsylvania; Department of Anesthesiology, Perioperative and Pain Medicine, Brigham and Women’s Hospital; Department of Biomedical Informatics, Vanderbilt University Medical Center; Department of Medicine, Vanderbilt University Medical Center; Department of Psychiatry and Behavioral Sciences, Vanderbilt University Medical Center; Center for Precision Psychiatry, Department of Psychiatry, Massachusetts General Hospital; Geisinger Clinic, Geisinger; Department of Imaging Science and Innovation, Geisinger; Neuroscience Institute, Geisinger; Department of Basic Sciences, Geisinger Commonwealth School of Medicine

**Author notes:** **Co-corresponding Authors:** Vanessa Troiani, Sandra Sanchez-Roige.

**Keywords:** opioids, prescription data, substance use disorders, electronic health records

## Abstract

**Background:** Electronic health records (EHR) are useful tools for understanding complex medical phenotypes, but they have been underutilized for opioid use disorders (OUD). Patterns of prescription opioid use might provide an objective measure of OUD risk.

**Methods:** We extracted data for over 2.6 million patients across three health registries (Vanderbilt University Medical Center, Mass General Brigham, Geisinger) between 2005 and 2018. We defined three groups based on levels of opioid exposure: No Prescription, Minimal Exposure (2 prescriptions within 90 days at least once, but never 3 prescriptions <90 days apart), and Chronic Exposure (≥10 opioid prescriptions in a year), and compared them to the full registries and to patients with OUD diagnostic codes. We extracted demographic and clinical characteristics known to co-occur with OUD, including psychiatric and substance use disorders, pain-related diagnoses, HIV, and hepatitis C.

**Results:** The prevalence of substance (alcohol, tobacco, cannabis) use disorders was higher in patients with OUD and Chronic Exposure than those with No Prescription or Minimal Exposure. Patients in the OUD and Chronic Exposure groups had more psychiatric (anxiety, depression, schizophrenia, bipolar disorder) and medical comorbidities (pain, hepatitis C, HIV) than those in the Minimal Exposure group. Notably, patients in the Minimal Exposure group had different comorbidity profiles (higher rates of substance use and psychiatric disorders, more pain conditions) than those in the Unscreened or No Prescription groups, highlighting the value of including opioid exposure in studies of OUD.

**Conclusions:** Long-term opioid prescription use may serve as an additional tool to characterize OUD risk.

## 1. INTRODUCTION

The opioid epidemic is a significant public health challenge in the United States, with continued high rates of hospitalizations and mortality as a result of misuse and abuse of prescription or illicit opioids (Gostin et al., 2017; Shipton et al., 2018). Opioid use disorder (**OUD**) evolves from a series of opioid consumption transitions, starting with exposure and continuing through regular use, misuse, abuse, dependence, and relapse (Kaye et al., 2017; Strang et al., 2020). Prevalence estimates for these phenotypes vary widely, in part due to variation in ascertaining and defining them (Freda et al., 2021). Indeed, one key challenge to defining opioid use phenotypes is the need to differentiate individuals across this spectrum of overlapping features.

Electronic health records (**EHR**) offer novel solutions for capturing opioid use behaviors in real-world healthcare settings, as they contain rich medical data relevant to OUD. Most OUD case definitions rely on diagnostic codes, but this approach is problematic, as OUD tends to be underdiagnosed (Guy and Zhang, 2018). Several opioid phenotype definitions have been developed to date that extend beyond diagnostic codes to include other sources of data, including prescription data available in the EHR (see **Supplementary Material**; Brummett et al., 2017; Butler et al., 2007; Calcaterra et al., 2018; Canan et al., 2017; Cochran et al., 2014; Coyne et al., 2021; Hylan et al., 2015; Karhade et al., 2019; Knisely et al., 2008; Sun et al., 2016; Webster, 2017; Webster and Webster, 2005).

However, little is known about patterns and correlates associated with different levels of prescription opioid exposure, including which factors distinguish patients across clinically distinct categories of exposure, and whether these patterns are consistent across different health systems. A better understanding of opioid use phenotypes and comorbidities across different levels of opioid exposure is beneficial in various data-driven research, including clinical prediction, treatment outcomes, diagnosis, prevention, epidemiology, and genomics. The work described here furthers our understanding of the clinical correlates of OUD and opioid use behaviors across three health systems, as part of the PsycheMERGE consortium. PsycheMERGE, which is an extension of eMERGE (McCarty et al., 2011), leverages EHR and genomic data for mental health research (Zheutlin et al., 2019), including substance use disorders (**SUD**).

In the present study, we characterized three opioid risk groups based on patterns of prescription opioid use and a fourth group based on International Classification of Diseases (ICD) diagnostic codes for OUD. Using data from three large health systems, we sought to: 1) evaluate and compare demographics and psychiatric and medical comorbidities across the four groups; 2) assess how the four groups differ in comparison with patients with no prescription data, and the general population of patients from each system; and 3) compare consistencies and differences in results across the three healthcare systems.

## 2. METHODS

### 2.1 Data Sources

Our data sources spanned three health systems: Vanderbilt University Medical Center (VUMC), Mass General Brigham (MGB), and Geisinger. Details of each registry, including demographics and data sources, are listed in the Appendix in the Supplement. We acquired Institutional Review Board approval (VUMC: 201767, MGB: 2018P002642); consent was not required for review of deidentified medical records. The Geisinger Institutional Review Board deemed this research exempt because all variables were extracted and summarized using an approved data broker.

Patients were included in the analyses if they had at least three years of medical history available between 2005 and 2018 and were 18 years of age or older on 12-31-2018. A minimum of three years of medical history was chosen to increase the likelihood that patients had enough prescription data to detect opioid prescription patterns. Patients younger than 18 years were excluded to reduce the likelihood of including individuals who had not yet developed OUD. We excluded patients with a cancer diagnosis (**Supplementary Table 1**) due to potential for long-term analgesia for cancer-related pain. We extracted relevant ordered (VUMC, MGB, Geisinger) or filled (Geisinger) opioid prescriptions using a list of commonly prescribed opioids (**Supplementary Table 2**). There were 627,396 patients from VUMC, 1,272,880 patients from MGB, and 733,637 patients from Geisinger who met the inclusion and exclusion criteria.

### 2.2 Prescription opioid phenotyping and group definitions

Five groups were included in the study (**Table 1**). First, we identified all patients from each health system who met inclusion/exclusion criteria (described above), which we refer to as the “Unscreened” group (aka the overall study sample). From this group, we next defined three sub-groups using inpatient and outpatient medication records based on prescription opioid exposure levels, derived from previously published work (Calcaterra et al., 2018; Sun et al., 2016). Patients in the “No Prescription” group had no documented opioid prescriptions during the period of observation. Patients in the “Minimal Exposure” group received two opioid prescriptions within 90 days at least once and no additional prescriptions within 9 months, but did not have 3 or more prescriptions no more than 90 days apart at any point during the period of observation (Katzman et al., 2020). Patients in the “Chronic Exposure” group received 10 or more opioid prescriptions within a 12-month period (Calcaterra et al., 2018; Sun et al., 2016). The final “OUD’’ group included patients with at least one ICD code for OUD (**Supplementary Table 3**). The definition of this group did not incorporate prescription data; therefore, patients in this group could overlap with the three prescription-based groups (code used to determine group membership available here: https://github.com/sanchezroigelab/OUD_spectrum_PsycheMERGE). The No Prescription, Minimal Exposure, Chronic Exposure and OUD groups cover only a subset of the patients in the Unscreened group.

**Table 1.**
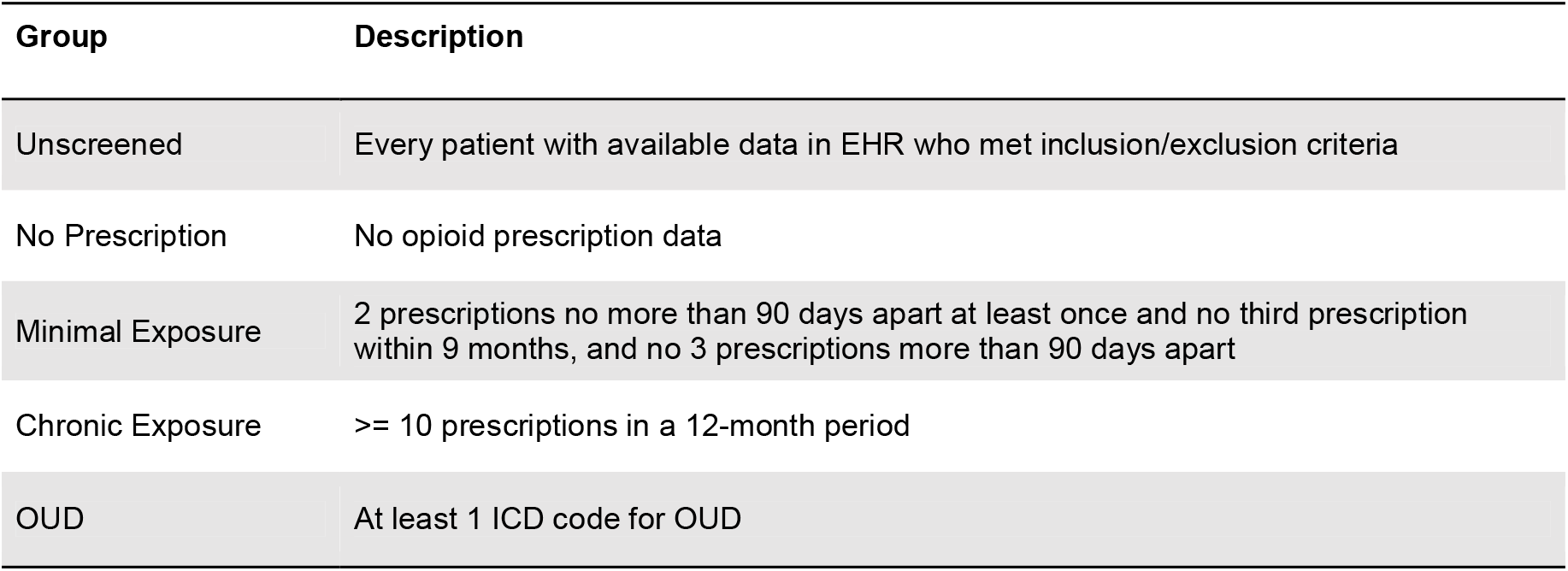
Description of each group used in the analyses. Only patients >= 18 years of age, with no history of cancer, and over 3 years of medical record history were included in the analyses. We avoid referring to the patients in the “No Prescription” group as having “no exposure” because exposure status is defined with prescription data and not verified by patient self-report.

### 2.3 Outcome Measures

Within each group, we characterized the length and density of EHR, demographics, opioid use patterns, and OUD diagnoses (**Supplementary Material**). Density was defined as the total number of non-unique ICD codes a patient received between 2005-2018 divided by the number of years included in the analysis. We also identified diagnoses previously identified as comorbid with OUD, including other SUD (Jones, 2019), psychiatric disorders (Jones, 2019), and other medical conditions, including HIV, hepatitis C, and pain-related diagnoses (Volkow et al., 2019) relevant ICD codes in **Supplementary Tables 4-16**).

To further examine patterns of prescription opioid use, we defined periods of exposure or “bouts” as at least two opioid prescriptions that occurred no more than 90 days apart. A bout was considered to end when there were more than 90 days between opioid prescriptions for a particular patient. We calculated the average number and length (in days) of bouts.

### 2.4 Statistical analyses

Descriptive statistics (frequency, percent, mean [M], standard deviation [SD]) were used to describe and qualitatively compare the different groups (**Supplementary Table 17**). Throughout the text, we present ranges (e.g., in percentages) across the three registries.

Demographic characteristics and outcomes across the three prescription-based groups were compared using Chi-square tests for categorical outcomes, and independent t-tests or Kruskal-Wallis tests for continuous outcomes. We regarded both a *p*<0.05 and a 5% difference in prevalence between any of the three prescription opioid groups clinically important. Given the large sample sizes, with only a few exceptions, outcomes were statistically different across groups and therefore only qualitative descriptions are provided in the results section. Full statistical results are described in **Supplementary Table 18**.

## 3. RESULTS

### 3.1 Demographics

The demographic composition for age and sex was similar across the Unscreened groups from different health system registries (**Supplementary Table 17**). Average age at the time of the analysis was 50.8-53.3 years, with 23.1-27.0% patients under age 35, and 41.3-44.6% male patients. Race/ethnicity varied within the registries, reflecting the geographic regions from which each health system draws. Yet the majority of the patients were identified in the EHR as White (74.1-95.4%), while only 3.2-12.6% were identified as Black or African American, 2.3-7.5% as Hispanic, 0.6-4.5% as Asian, and 0.4-9.0% as other race/ethnicity.

A higher proportion of patients in the OUD group were male (44.8-60.7%) compared to the prescription-based groups (30.9-46.2%); this sex difference was largest in the MGB registry (**Supplementary Table 17**). Patients in the OUD group were 5-8 years younger, on average, than the Unscreened group, and 12-15 years younger than patients in the Chronic Exposure group. Only 5.7-7.6% of the patients in the Chronic Exposure group were under the age of 35, compared to 15.3-33.7% for the OUD, No Prescription, and Minimal Exposure groups. The Chronic Exposure and OUD groups were predominantly composed of patients identified as Whites (81.7-96.8%) and Blacks/African Americans (2.3-13.9%), with lower proportions of Hispanics (1.0-5.0%) and Asians (0.1-0.9%) in comparison to the other groups (71.1-96.5% Whites, 2.7-16.0% Black/African Americans, 0.3-7.4% Hispanics, and 0.1-1.1% Asians).

Length of EHR was greater for the Minimal Exposure (11.0-13.9 years), Chronic Exposure (11.4-14.6 years) and OUD (11.7-13.0 years) groups compared to patients in the Unscreened (9.9-11.9 years) and No Prescription (9.3-11.0 years) groups. Density of EHR was highest for the Chronic Exposure group (24.9-59.3), followed by the OUD group (15.5-33.1).

### 3.2 Prevalence of OUD diagnoses

The overall prevalence of an OUD diagnosis in the Unscreened group was 1.2-1.7% (Figure 1); this number fluctuated during the study period (**Supplementary Figure 1**), with a steeper increase around 2013 in VUMC and MGB and a later peak (2017) in Geisinger.

**Figure 1.**
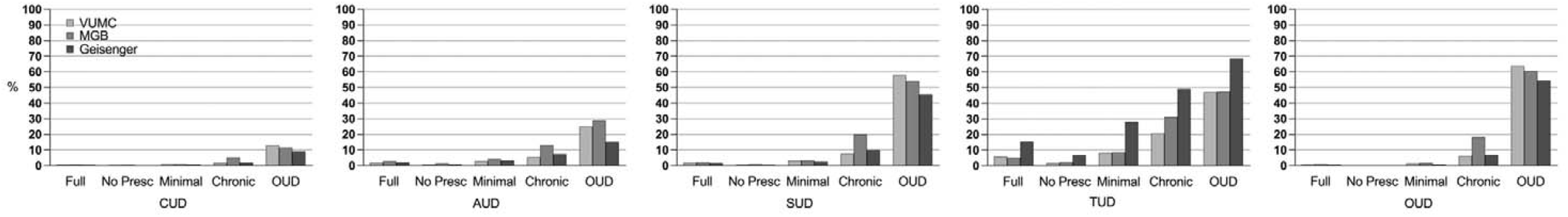
Rates of substance use disorders across the three registries (VUMC, MGB, Geisinger) by levels of opioid exposure (Unscreened, N[range across registries]=627,396-1,272,880; No Prescription, N= 251,546-582,542; Minimal Exposure, N= 50,112-70,510; Chronic Exposure, N= 14,373-27,507; OUD, N= 8,673-21,489). *Full*, Unscreened group; *No Presc*, No Prescription group; *CUD*, cannabis use disorders; *AUD*, alcohol use disorders, *SUD*, substance use disorders; *TUD*, tobacco use disorders; *OUD*, opioid use disorders. Note that OUD as outcome pertains to having two or more OUD ICD codes on separate occasions.

OUD diagnoses were higher in non-Hispanic Whites and increased with exposure to prescription opioids - from 0.3-0.6% for No Prescription, to 1.6-2.7% for Minimal Exposure, and 9.0-24.4% for Chronic Exposure. These patterns were consistent across health systems. 82.1-96.8% of the patients in the OUD group were non-Hispanic Whites, which was higher than other racial/ethnic groups (0.1-10.4%).

### 3.3 Opioid prescription patterns

Average age at first opioid prescription ranged from 45.7 to 46.2 years in the Unscreened group across health systems, with the youngest average age of first prescription observed in the OUD group (34.8-39.0) and oldest among the Chronic Exposure group (46.8-52.5).

The duration and number of periods of opioid exposure (“bouts”) increased as exposure to prescription opioids increased, with the Chronic Exposure group having the highest number (3.3-4.1) and length (224.1-566.5 days) of bouts. The OUD group had the second highest number (1.4-3.0) and length of bouts (135.8-185.0 days). In the Unscreened and Minimal Exposure groups, patterns of use diverged. For example, in the Unscreened group, the average number of bouts ranged from 0.4-1.8, and length of bouts ranged from 12.6-97.0 days. In contrast, the Minimal Exposure group had a higher number of bouts (1.5-1.8) but the length of use was shorter (10.0-15.9 days).

### 3.4 Substance use disorders

The prevalence of SUDs was highest in the OUD group (alcohol: 15.2-28.8%, tobacco: 47.0-68.6%, cannabis: 9.1-13.0%; Figure 1). In contrast, the No Prescription group showed dramatically lower rates of SUDs (alcohol: 0.9-1.5%, tobacco: 2.3-6.8%, cannabis: 0.2-0.5%) than any other group, including the Minimal Exposure (alcohol: 3.1-4.1%, tobacco: 8.6-28.1%, cannabis: 0.8-1.2%) and the Chronic Exposure (alcohol: 6.1-13.2%, tobacco: 26.6-49.2%, cannabis: 1.9-5.1%) groups.

### 3.5 Psychiatric disorders

Prevalence of psychiatric disorders was highest in the Chronic Exposure and OUD groups (Figure 2). For example, the prevalence of anxiety, one of the most common psychiatric disorders observed, was 19.3-37.2% and 25.7-36.6%, respectively, for the Chronic Exposure and OUD groups, compared to 8.0-17.3% for the Minimal Exposure group, 4.2-7.2% for the No Prescription group, and 6.5-11.5% for the Unscreened group. Depression prevalence was higher (36.4-50.4%) in the Chronic Exposure and OUD groups compared to the other groups (6.3-20.0%).

**Figure 2.**
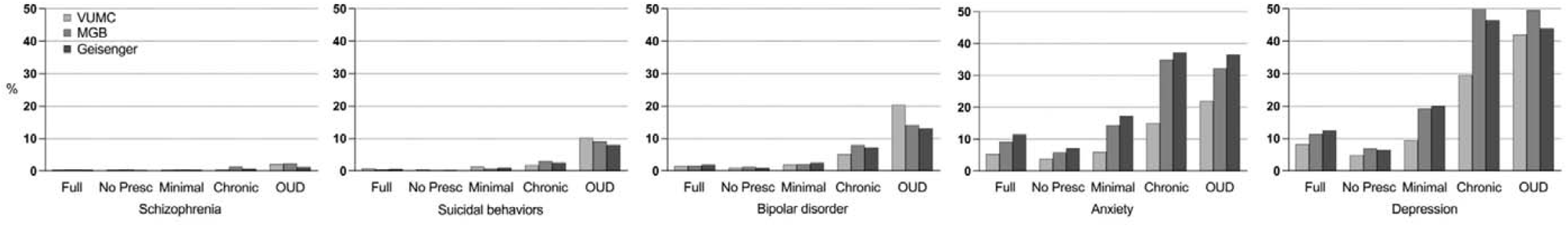
Rates of psychiatric disorders across the three registries (VUMC, MGB, Geisinger) and levels of opioid exposure (Unscreened, No Prescription, Minimal Exposure, Chronic Exposure, and OUD). *Full*, Unscreened group; *No Presc*, No Prescription group.

Bipolar disorder prevalence was highest in the OUD group (13.1-21.1%), higher than the Chronic Exposure group (5.2-8.0%), and dramatically higher than the Minimal Exposure (2.0-2.6%), No Prescription (1.0-1.3%) and Unscreened (1.6-1.9%) groups. Schizophrenia prevalence in the OUD group was slightly elevated (1.2-2.4%) compared to all other groups.

Suicidal behavior prevalence was only higher in the OUD group (8.0-11.5%), compared to all other groups (0.2-3.1%).

### 3.6 Pain and other medical conditions

As expected, a large percentage of patients exposed to opioids had pain conditions (Minimal Exposure: 63.9-76.0%; Chronic Exposure: 92.2-96.6%; Figure 3), more so than the No Prescription or Unscreened groups (30.8-33.9% and 48.8-51.6%, respectively).

**Figure 3.**
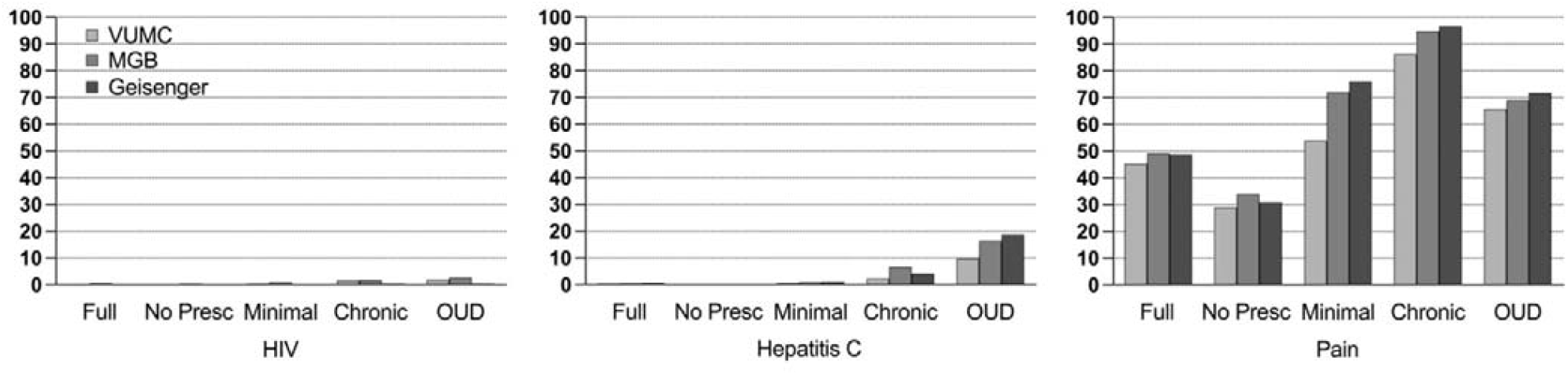
Rates of medical conditions (HIV, hepatitis C, pain) known to be comorbid with opioid use disorders across the three registries (VUMC, MGB, Geisinger) and levels of opioid exposure (Unscreened, No Prescription, Minimal Exposure, Chronic Exposure, and OUD diagnosis). *Full*, Unscreened group; *No Presc*, No Prescription group.

The prevalence of HIV and hepatitis C was highest in the Chronic Exposure and OUD groups, particularly for hepatitis C (2.6-6.7% and 9.7-18.6%, respectively), compared to the other groups (0.1-1.2%).

## 4. DISCUSSION

Detecting OUD in EHR analyses is notoriously challenging. Diagnostic codes can be insufficient because OUD tends to be underdiagnosed (Hallgren et al., 2021; Kirson et al., 2015). Furthermore, models often focus on extreme opioid use in service of predicting case/control classifications and thus miss the full spectrum of opioid use behaviors (Palumbo et al., 2020). Our approach overcame these limitations by examining a continuum of opioid use behaviors. This approach is particularly important for opioid use phenotypes because behaviors range from appropriate medical use of prescribed opioids to abuse of prescription and illegal opioids.

Our study is the first to systematically identify patterns and correlates of short and long-term prescription opioid use and OUD across health systems, which facilitated important observations. First, we found that the OUD group had unique characteristics compared to the other groups, the most salient of which included comorbid psychiatric (anxiety, depression) and substance use disorders, particularly tobacco use disorders, in line with previous findings (Barry et al., 2016; Edlund et al., 2010; Nazarian et al., 2021; Volkow et al., 2019). Prior studies estimated that 45-57% of individuals with OUD had at least one psychiatric disorder and reported that polysubstance abuse was exceedingly common (Freda et al., 2021), comparable to our findings. In addition, average age at first opioid prescription among the OUD group was approximately 10 years younger than that of other groups, highlighting the importance of age at first exposure to prescribed opioids and onset of OUD (Phillips et al., 2017). The demographic factor most noticeably associated with the OUD group was EHR-identification as Non-Hispanic White. This finding is consistent with previous literature suggesting that the opioid epidemic in the United States has historically primarily affected rural and suburban Non-Hispanic Whites (CDC, 2019; Keyes et al., 2014).

Second, our findings emphasize the value of including opioid prescriptions in assessing risk for OUD (Rentsch et al., 2019; Naumann et al., 2019). For example, with the exception of age at first opioid prescription, patients in the Chronic Exposure group most closely resembled the OUD group across most of the characteristics evaluated and may therefore represent the group with highest risk of having or developing OUD (Klimas et al., 2019; Volkow et al., 2019). Consistent with this finding, the Chronic Exposure group also had a higher rate of OUD diagnosis than the No Prescription or Minimal Exposure groups. In addition, OUD diagnoses increased from the No Prescription to the Minimal Exposure group, providing further evidence for the relevance of assessing opioid prescriptions when determining risk for OUD (Vowles et al., 2015).

Although findings were generally consistent across sites, we did observe some heterogeneity. This may have been due to, in part, differences in underlying patient populations, as evidenced by the differences in race/ethnicity proportions in the Unscreened groups; prior studies have documented varying correlates of prescription opioid misuse by race/ethnicity (Nicholson, 2018). Differences across sites can also be due to a more rural population at Geisinger compared to MGB and VUMC (Buettner-Schmidt et al., 2019) and the challenges of access to behavioral health services and treatment in serving rural populations. Nonetheless, the overall consistency in findings suggests that correlates of opioid use phenotypes are shared between healthcare systems despite differences in data recording and patient populations, and that opioid prescription EHR-based studies from different systems can be compared to one another, if using similar definitions.

Our findings have relevance for future EHR-based OUD research. First, our work re-emphasizes the importance of incorporating opioid prescriptions when defining the spectrum of opioid misuse and OUD. Prior algorithms have incorporated opioid prescriptions to identify opioid misuse (e.g., Calcaterra et al., 2018; Canan et al., 2017; Rough et al., 2019), but additional efforts could place patients on a spectrum of problematic opioid use behaviors.

For example, to identify individuals at risk for developing OUD, phenotype risk scores (Bastarache et al., 2019; Ruderfer et al., 2020) could be constructed by agnostically training and testing a risk model using diagnosis codes (for OUD and other relevant predictors) and prescriptions. Such a strategy would not only acknowledge OUD risk in the absence of an OUD diagnosis, but could also allow for modeling of OUD risk trajectories over time (Elmer et al., 2019). Second, our descriptive data confirms the relevance of several predictors of opioid misuse (e.g., age, substance and psychiatric comorbidities) that have been previously incorporated (but not necessarily validated; Canan et al., 2017; Schirle et al., 2021) into algorithms identifying opioid misuse. Third, the depth and breadth of data in EHR registries can be leveraged to clarify the phenotypic structure of OUD phenotypes. Future studies could use data-driven methods to integrate additional EHR components (e.g., prescriptions for other controlled substances, types of pain diagnoses, opioid dosages and types) and implement cluster-based methods such as latent profile analysis, k-means clustering, or principal component analysis to explore OUD sub-phenotypes (Nylund et al., 2007).

Lastly, our work has implications for genetic studies, in particular informing selection of individuals for genetic analyses (Sanchez-Roige & Palmer, 2020). A major roadblock in conducting genome-wide association studies (GWAS) of opioid use phenotypes is the lack of opioid exposed controls, resulting in the frequent use of unscreened individuals as controls. Consistent with previous work showing that using unscreened controls can introduce biases in genetic analyses (Polimanti et al., 2020), our work demonstrates that a Minimal Exposure group has a different set of clinical characteristics than an Unscreened or No Prescription group.

### 4.1 Limitations

This study is subject to several limitations. We lacked information about the reasons for prescribing opioids, which could help differentiate patients with problematic opioid use. Similarly, we did not have complete information on opioid dosages (Morasco et al., 2010) (and therefore, morphine milligram equivalents) or information on use of illicit opioids, and we were not able to differentiate between opioid types, which would have helped identify additional misuse phenotypes, such as rapid dose escalation trajectories (Rentsch et al., 2019). Further, we relied on opioid prescriptions to index opioid use, but this is likely an imperfect proxy for actual use. In addition, we did not include buprenorphine as a qualifying opioid for the opioid prescription groups due to its use in treating OUD; a similar case could be made for methadone, but we included it because it is often used to treat pain. These decisions could have led to misclassification of patients. Reliance on OUD diagnosis for the OUD group could also have led to misclassification given the underdiagnosis of OUD (Palumbo et al., 2020); this may have been particularly problematic in earlier years included in the study. Furthermore, the associations observed represent correlations and not causation; future studies may include sequences of events to disentangle potential trajectories of effect between the groups and the outcomes. Lastly, it is not known whether these results are generalizable to other populations, but the external validity of our findings is supported by consistencies observed across the three health systems, which serve diverse patient populations.

## 5. CONCLUSION

This work can inform the selection of cases and controls for epidemiologic and genetic studies, demonstrates the utility of using levels of prescription opioid use in elucidating different aspects of OUD pathophysiology, and supports the appropriateness for future meta-analyses across health systems.

## Data Availability

Data produced in the present study are available to researchers at the cited institutions. Code used to determine group membership available here: https://github.com/sanchezroigelab/OUD_spectrum_PsycheMERGE

## ACKNOWLEDGEMENTS AND FUNDING

MVJ, SBB and SSR were supported by funds from the California Tobacco-Related Disease Research Program (TRDRP; Grant Number T29KT0526), SBB is also supported by P50DA037844, SSR is also supported by NIDA DP1DA054394. HL is supported by funds from NIH 1R01MH118233-01. BJC is supported by funds from NIMH R01 MH121924. ABF was supported by the Training Program On Genetic Variation And Human Phenotypes Training Grant (T32GM080178). MP is supported by funds from NIDA K01DA04993. TTM is supported by funds from NIH T32HG010464. CGW is supported by funds from NIMH R01 MH118233, R01 MH121455, R01 MH120122, R01 MH116269, DoD MSRC W81XWH-10-2-0181, and the FDA WO2006. VT and RCC are supported by funds from NIDA R01DA044015 and a grant from the Pennsylvania Department of Health. The PsycheMERGE consortium is supported by NIMH R01 MH118233 (JWS, LKD).

The project using VUMC data was supported by CTSA award No. UL1TR000445 from the National Center for Advancing Translational Sciences. Its contents are solely the responsibility of the authors and do not necessarily represent official views of the National Center for Advancing Translational Sciences or the National Institutes of Health.

The content is solely the responsibility of the authors and does not necessarily represent the official views of the National Institutes of Health.

