## Supplementary Material for "Identifying high-risk comorbidities of short and long-term opioid prescription use"

PsycheMERGE Opioid Use Disorder Workgroup

**Supplementary Figure 1**

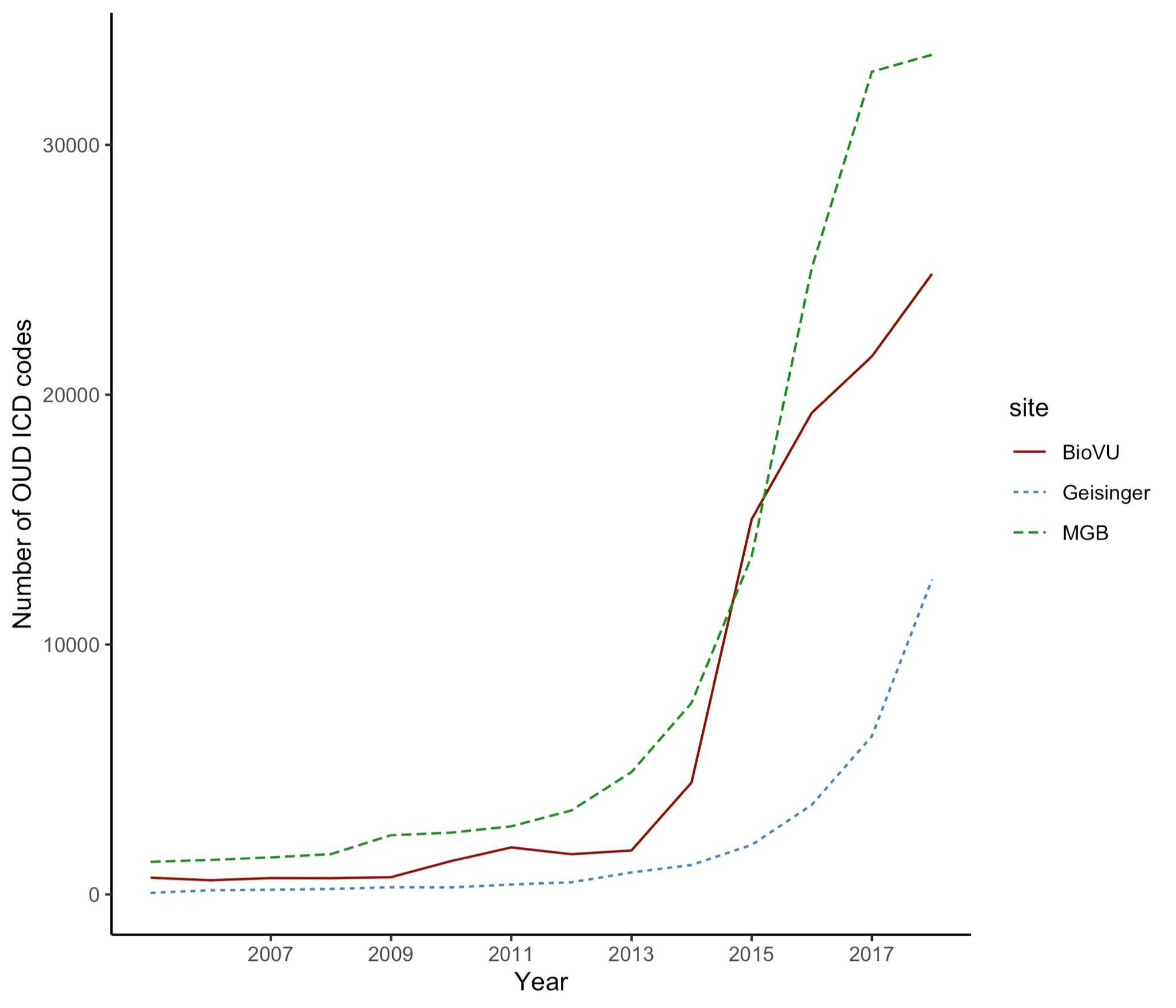

Overall prevalence of an OUD diagnosis across the three health systems (VUMC, MGB, Geisinger).

**METHODS**

**Phenotyping definitions to capture opioid behaviors**

Below we describe the most common definitions to capture various opioid behaviors. Phenotypic descriptions range from clinical diagnosis to characterization of novel opioid phenotypes, such as new persistent use and dose escalation, to data driven phenotypes utilizing artificial intelligence and machine learning. Phenotypes surrounding opioid use, misuse, abuse, and addiction have been defined using different data sources, including diagnostic codes, prescription databases and/or insurance claims, and clinical notes.

**International classification of disease (ICD) codes to capture OUD.** One of the most commonly used coding systems in the EHR include ICD codes, which comprise a standardized medical classification system utilized for disease diagnosis, injury, and cause of death. The use of ICD codes for definitions of OUD can include both prescription opioid use and use of illicit opioids. Individuals lacking OUD ICD codes often serve as controls, which can contain a mixture of exposed and unexposed individuals.

**Prescription data for a longitudinal characterization of opioid use patterns.** Prescription data can be used to derive a variety of sophisticated phenotypes, including new persistent use following surgery, chronic levels of use, and escalation of use. For example, approximately 6% of opiate naive patients who undergo surgery will continue to use opioids more than three months after their surgery; this phenomenon is termed “new persistent opioid use” (Brummett et al., 2017). This phenotype can be derived using a variety of definitions that vary in terms of how restrictive they are, including the need to access national prescribing data. Definitions differ in terms of the number of prescriptions and set time period for which the patient must fill opioid prescriptions. Chronic prescription opioid use has been previously defined as having filled 10 or more opioid prescriptions over 1 year (e.g. (Calcaterra et al., 2018; Sun et al., 2016)). Another phenotype that has utilized longitudinal prescription data characterizes patterns of opioid prescribing, or escalation of use, by modeling trajectories based on morphine equivalent daily dose (Wei et al., 2019). Both new persistent use and escalation of dose require assurance of complete/comprehensive prescription data, which is often not available across sites. Instead, we compiled a chronic use definition, which can be implemented across sites and is more tolerant to missingness. Prescription data can also be used to define exposed controls. Exposed controls are individuals who have been exposed to opioids medically or illegally who did not later develop OUD (Polimanti et al., 2020).

**Questionnaires and clinical notes to define opioid use, misuse, OUD.** Opioid use behaviors can be examined via questionnaires. These are often used in clinical context, although the data availability is often restricted by the need of using natural language processing (NLP) technologies. Questionnaires utilized to define opioid use and misuse include the Opioid Risk Tool (Webster and Webster, 2005), the Current Opioid Misuse Measure (Butler et al., 2007), the Prescription Opioid Misuse and Abuse Questionnaire (Coyne et al., 2021), the Prescription Opioid Misuse Index (Knisely et al., 2008), and the Addiction Severity Index (McLellan and et al., 1980). Characterization of predictive factors for continuous use has also been carried out via automable algorithms (Cochran et al., 2014; Karhade et al., 2019). Such algorithms can facilitate identification of patients in need of intensive screening or intervention for nonmedical opioid use (Canan et al., 2017). Lastly, NLP tools have been utilized to search the free text clinical notes of the EHR to facilitate identification of problematic opioid use phenotypes (Hylan et al., 2015). Pre-existing clinical NLP techniques include MedLEE (Medical Language Extraction and Encoding System), SymText, KnowledgeMap, LifeCode, FreePharma, and Coderyte (Xu et al., 2010). While these methods have proven sufficient to capture opioid behaviors, limitations exist when processing text with multiple signatures or highly contextual information (Xu et al., 2010).

**Data sources**

VUMC is a tertiary care center that provides inpatient and outpatient care in Nashville, Tennessee. The VUMC EHR was established in 1990 and contains data on more than 3.2 million individuals. The population was 56% female, with approximately 81% of patients being identified in the EHR as Caucasian, 13% African American, 2% Asian, 3% Hispanic, and 4% other races and ethnicities. The protocol was approved by the VUMC Institutional Review Board (IRB #201767). Medication data was extracted using a natural language processing system (MedEx), which extracts medication information from clinical notes (Xu et al., 2010).

MGB Research Patient Data Registry (RPDR; <https://rpdrssl.partners.org/>) is an EHR database which spans more than 20 years of data from over 6.5 million patients seen in Massachusetts General Hospital, Brigham and Women’s Hospital, and other community and specialty hospitals in the Boston area. The population was 56% female, with approximately 75% of patients identified in the EHR as Caucasian, 7% Black or African American, 5% Asian, 7% Hispanic, and 10% other races and ethnicities. Medication data were derived from several source systems including Longitudinal Medical Record (LMR) from MGB affiliated hospital pharmacies, TSI, OnCall, EPIC, EPSi, PAML, and Paragon.

Geisinger is an integrated health system with its own health plan that serves over 3 million Pennsylvania residents within an area with low outmigration, leading to a long duration of patient records (average ~20 years). Geisinger implemented the EPIC EHR in 1996. The population was 57% female, with approximately 91% of patients identified in the EHR as non-Hispanic White, 5% Hispanic, 3% African American, and 1.5% other (including Asian, Pacific Islander, American Indian, or more than one race/ethnicity). Medication data includes those prescribed and dispensed by Geisinger providers at Geisinger pharmacies and national prescribing data obtained from Surescripts.

**Outcome measures**

Demographic variables included gender, age, percentage of patients under 35 years old, EHR-reported race, length of record and density of record.

Opioid metrics included incidence of OUD diagnosis, and patterns of prescription opioid usage. Precisely, we evaluated the prevalence of patients with ICD codes for OUD, average age at first opioid prescription, average age at first OUD ICD diagnosis (in years), and incidence of opioid overdose across the different groups.

Substance use disorders are highly comorbid in individuals diagnosed with OUD (Webster, 2017). To examine other substance use disorders, including alcohol use disorder, tobacco use disorder, and other substance abuse, we calculated the percentage of patients with each diagnosis across the groups.

Similarly, a large proportion of OUD patients have at least one other comorbid psychiatric disorder (Barry et al., 2016; Brooner, 1997; Kidorf et al., 2004; Sullivan et al., 2006). We examined the psychiatric disorders that were most common in the registries and that are known to co-occur with OUD based on prior literature. In particular, we examined the rates of anxiety, depression, suicidal behavior, schizophrenia and bipolar disorder across the groups.

Experience of pain increases the prevalence for prescription opioid use and opioid misuse vulnerability (Barth et al., 2013; Nazarian et al., 2021). For each definition, we calculated the percentage of patients with pain conditions as defined via ICD, the average number of pain codes and the average number of different pain conditions. We also examined the percentage of patients who had other medical conditions associated with pain, such as hepatitis C and HIV. Frequently, OUDs are comorbid with infectious diseases because injecting drugs increases the risk of blood-transmitted infections, including HIV and hepatitis C virus infections.

All outcome measures were defined as the presence of at least 2 ICD codes on separate occasions. Complete lists of ICD codes are available in the Supplementary Tables.
